# Responsiveness of Health-Related Field-Based Physical Fitness Tests in Adults: The Adult-Fit Project

**DOI:** 10.1101/2025.07.12.25331202

**Authors:** Magdalena Cuenca-Garcia, Carolina Cruz-León, José Jiménez-Iglesias, Sandra Sánchez-Parente, Víctor Segura-Jiménez, Francisco B. Ortega, José Castro-Piñero

## Abstract

The aim of the present study was to determine the responsiveness of the health-related field-based physical fitness tests used in adults. A total of 62 non-active participants aged 18 to 64 years were randomized into the intervention (*n*=31) or control group (*n*=31). The exercise program included 3 sessions/wk (60 min/session) of multicomponent exercise training for 12 weeks. The control group continued with their usual routines. Pre-post differences were explored with pairwise comparison ANOVA for each group. Percentage of change and the size effect were also calculated. The proportion of responders/non-responders and percentage of the population that was expected to respond to the intervention were calculated. Characteristics of study groups were similar at baseline (all *P*>0.05). All tests were found to be responsive (all *P*<0.01) after the exercise program, except the waist circumference measurement (*P*=0.09); with an effects size of moderate to large (*Cohen’s d* >0.50), except to the weight, body mass index, waist circumference and handgrip tests with trivial effect sizes (*P*>0.05, *Cohen’s d*<0.20). The highest proportion of individual responders based on the standard deviation was observed in the 4×10-m shuttle run, 30-s sit-to-stand, 6-m gait speed and timed up & go tests. The proportion of the population that was expected to respond to the intervention was higher than 85% in most of the field-based physical fitness tests evaluated. Overall, all health-related field-based physical fitness tests were found to be responsive after a multicomponent exercise intervention regardless sex and age.

## Introduction

Physical fitness is a powerful predictor of morbidity and mortality, and its assessment is therefore a useful indicator for public health monitoring [1–4]. However, physical fitness assessments are not routinely used in clinical settings to assess or predict health, in part due to the use of laboratory testing, which is an objective and accurate method of assessing physical fitness that requires sophisticated equipment. Additionally, the ratio administration time-evaluator-participant are not efficient in clinical settings.

Field-based physical fitness tests could be an excellent alternative, in this line. The aim of the ADULT-FIT study, supported by the Spain Ministry of Economy, Industry and Competitiveness, was to propose a health-related field-based physical fitness test battery based on their criterion-validity (i.e., output of the test correlates with the criterion measure, e.g., the gold standard) [5, 6], predictive validity (i.e., relationship with health outcomes) [4, 7, 8], reliability (i.e., reproducibility of values in repeated trials on the same individual) [9], feasibility (i.e., degree of being conveniently done) [10], safety (i.e., number of health complications occurring during the testing procedure) [10], and responsiveness or longitudinal validity (i.e., ability of a test to detect changes over time) to be used in adults [11]. It is important that the health-related field-based physical fitness test battery should meet the criteria quality mentioned above to an efficient and appropriate use. We have previously studied the validity, reliability, feasibility and safety of the health-related field-based physical fitness tests used in adults [5, 9, 10]. Unfortunately, the assessment of responsiveness is not usually considered when proposing a health-related field-based physical fitness test in health adults.

The responsiveness of a field-based physical fitness test is relevant in outcome studies as well as practical application in different settings (e.g.: clinical, sport club, etc). A field-based physical fitness test battery can be deemed to be valid, reliable, feasible and safe but if the test is unable to detect the relatively small changes in response to an intervention, it cannot be considered useful and appropriate to evaluate the effectiveness of an intervention. Thus, the aim of the present study was to determine the responsiveness of the health-related field-based physical fitness tests used in adult population.

## Methods

The present study is part of a national project: the ADULT-FIT study (REF: DEP2017-88043-R), whose main aim was to propose a health-related field-based physical fitness test battery in adults, based on their criterion-validity, predictive validity, reliability, feasibility, safety and responsiveness.

### Participants

Sample size was calculated according to the T-test between differences two independent mean (two groups) with a one-sided, a level of significance of 5%, an effect size of 0.80, a power of 90%, and considering a dropout rate of 10% (G*power 3.1.9.6, Düsseldorf, Germany). Participants were recruited through leaflets, local newspapers, and social media. A total of 62 non-active participants aged 18 to 64 years were recruited and randomized into the intervention group (*n*=31) or the control group (*n*=31). The inclusion criteria were: *(i)* being an adult (18-64 years old) and non-active; *(ii)* not having physical or mental illness that prevents from doing physical activity; *(iii)* have intention to carry out all the tests included in the study and; *(iv)* being able to read and understand the informed consent as well as the aim of the study. The exclusion criteria for this study were: *(i)* having acute or terminal illness; *(ii)* myocardial infarction three months before starting the study; *(iii)* unstable cardiovascular disease; *(iv)* medical prescription that prevents the performance of the tests and; *(v)* injury or circumstance that makes it impossible to carry out the tests correctly.

Participants were initially classified as active/non-active when following/not following World Health Organization recommendations for adults (https://www.who.int/). The following self-reported question was asked: how many days (in a typical week) do you practice physical activity/exercise or some sport, of at least moderate intensity, lasting at least 50 min per day?

All interested volunteers provided written informed consent to participate in the present study. The study was approved by the Committee for Research of Cádiz.

### Protocol Design

Physical fitness was assessed using the health-related field-based physical fitness tests at baseline (pre-test) and after 12-weeks (post-test) in all participants. The tests were carried out in three non-consecutive days, with at least 72 hours apart, in an indoor/outdoor facility under appropriate conditions (i.e., space, surface, equipment and sports clothing), and were assessed and supervised by researchers with experience in field-based physical fitness testing. The tests assessments were distributed in the following order: *i) 1^st^ day*: body weight, height, waist circumference, 4×10-m shuttle run, handgrip strength and 2-km walk tests; *ii) 2^nd^ day*: standing long jump, prone bridging and 6-min walk tests and; *iii) 3^rd^ day*: timed up & go, 6-m gait speed, 30-s sit-to-stand and 20-m shuttle run tests.

Post-test evaluation was conducted 72 hours after the conclusion of the last season of the intervention period under the same conditions, with researchers following the same protocol as in the pre-test evaluation.

Participants in the intervention group engaged in a 12-week multicomponent exercise; while those in the control group maintained their usual routine. Possible changes were then registered.

### Field-based Physical Fitness Measures

#### Body composition

Body weight (nearest 0.1 kg), height (nearest 0.1 cm) and waist circumference were measured following the protocol described by the International Society for the Advancement of Kynanthropometry [12]. Measurements were conducted sequentially on two occasions by the same rater (to account for intra-rater variability) and the mean between measurements was calculated. The raters were of the same sex as the participants. Body mass index (BMI) was calculated as weight (kg) divided by squared height (m^2^).

#### Cardiorespiratory fitness

The 20-m shuttle run, 2-km walk and 6-min walk tests were selected to measure cardiorespiratory fitness [13–16]. One attempt for each test was performed. Number of stages reached in the 20-m shuttle run test, total time employed to cover 2-km walking and total distance covered in 6-min walking were registered.

#### Motor fitness

The tests selected to measure motor fitness were the 6-m gait speed, timed up & go and 4×10-m shuttle run. In the 6-m gait speed test, one attempt was performed, and time employed to cover a distance of 6 meters walking as fast as possible was registered [17]. For the timed up & go test, one attempt was developed and total time employed in get up from the chair, walk 2.45-m (8 feet) as fast as possible, turn, and return to seated position was registered [16]. In the 4×10-m shuttle run test, two non-consecutive attempts were developed and the best attempt (minimum time spent in running and turning 4×10-m) was registered [18].

#### Muscular strength

Handgrip strength, standing long jump, 30-s sit-to-stand and prone bridging tests were selected to measure muscular strength. In the handgrip test, two non-consecutive attempts were developed with each hand and the best result at each grip was registered [18, 19]. For the standing long jump test, two non-consecutive attempts were performed and the best attempt (maximum distance reached with both feet together) was registered [18]. In the 30-s sit-to-stand test, one attempt was performed and sit down and stand up repetition in 30 s was registered [16]. For the prone bridging test, one attempt was performed and the total time in isometric position was registered [20].

### Training Program

The exercise program included 3 supervised sessions per week of multicomponent exercise (i.e., including aerobic, muscle strengthening and motor fitness activities) [21–23] for 12 weeks, based on the guidelines proposed by the American College of Sports Medicine for the prescription of physical exercise in healthy adult [24]. Each session lasted 60 minutes, 10 minutes of warm-up, 45-minutes of multicomponent exercise and 5 minutes of cool-down. Exercise intensity during sessions was checked individually by rating of perceived exertion on the Borg Scale (1-10 points) [25] and heart rate using the activity bracelet (Xiaomi Mi Band 4) [26]. The target was set between 5 and 8 on the Borg Scale (perceived “somewhat hard” or “hard” exertion) and between 60% and 91% of the reserve heart rate.

On one hand, the aim of the muscle strengthening and motor fitness exercises was to develop strength, stability and coordination and speed of movement. It was based on basic strength work aimed at fundamental motor patterns using free weights, elastic bands and self-loading. The exercises were: pulls and pushes (both horizontal and vertical) as well as hip/knee dominant exercises and lumbo-pelvic stabilization work. In terms of external load, the exercises progressed throughout the sessions in qualitative terms. They became increasingly complex and demanding from a motor point of view, by progressively including unilateral work, cross-chain and homolateral execution, combinations of patterns (e.g., hip dominance plus push-off) and more complex combinations (e.g., hip dominance plus push-off and hip dominant plus vertical thrust). The exercises also involved increasingly demanding bases of support from a stabilization point of view (e.g., transitioning from doing an exercise while standing to performing it in a split stance or on one leg). From a quantitative point of view (i.e., kilograms to be displaced), the participants were urged to mobilise increasingly heavier implements, thicker rubber bands and to adopt more demanding angles in the exercises with self-loads.

On the other hand, the aim of aerobic exercises was to develop cardiorespiratory fitness; for this purpose, intermittent work dynamics such as *High Intensity Interval Training* (HIIT), *As Many Repetitions as Possible (AMRP)* and *Fractional Running* methods were established. HIIT and AMRP consisted of alternating periods of high intensity (relative to the capacity of each participant) based on different types of movements (frontal running, lateral movements, jumps, etc.) and strength exercises with very light loads. The evolution of the external load involved accumulating progressively more effective time of advanced cardiovascular work, increasing from 12-15 to 30-35 min (modifying the total duration of the intensity periods, as along with their numbers). To enhance motivation and adherence, variations were introduced in both the exercises and training methods.

### Statistical Analysis

The distribution of each variable was examined with Kolmogorov-Smirnov normality test. All data were normally distributed. To compare the baseline variables between groups, the one-way ANOVA test was used.

To compare pre-post differences, we used pairwise comparison ANOVA for each group separately. Percentage of change (% Change) for each variable was calculated as: %Change = (mean (Post-test value) – mean (Pre-test value) / mean (Pre-test value)) * 100. *Coheńs d* was computed to quantify the magnitude of the effect size as standardized mean difference. The criteria to interpret the magnitude of the effect size were as follows: ˂0.2 trivial, 0.2-0.5 small, ˃0.5-0.8 moderate and ˃0.8 large [27]. Statistical analyses were performed using the Statistical Package for Social Sciences (IBM SPSS Statistics for Mac Os, version 29.0; Armonk, NY). Differences were considered significant when P <0.05.

Responder analysis was performed using a two-step approach: 1) pre-post change and an 80% confidence interval (CI) were calculated for each participant (80%CI classifies individuals with 90% certainty and it has been proposed as a default value) [28]. CI calculations were adjusted for sample size by using the following formula:

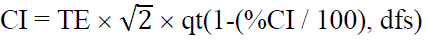

Where TE represents the typical error (i.e., standard deviation of the pre-post change divided by √2), qt() is the quantile function of the central t-distribution, %CI the desired width of the CI (80 in our analysis) and dfs the degrees of freedom (i.e., sample size – 1).

Then, the smallest worthwhile change (SWC) was calculated for every variable as 0.2 × the baseline between individual standard deviation. Finally, participants were classified as *likely responder* if the CI lies completely over or below the SWC depending of the interpretation of positive effect for each of the variables [29]. 2) We assume that each studied variable follows a normal distribution in the population with the mean observed score change and the standard deviation of individual responses (σ_IR_) as mean and standard deviation, respectively. σ_IR_ was calculated with the following formula:

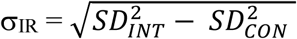

where 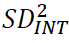 is the square of the calculated standard deviation of the observed score change from the intervention group, and 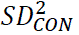 is the square of the calculated standard deviation of the observed change scores from the control group [30]. Next, the percentage of responders was calculated as the area of the normal distribution that lies beyond the SWC [31]. These analyses were performed with the R software environment for statistical computing [32].

## Results

Sixty participants finally completed the study (62% female); 31 and 29 in control and intervention group, respectively. The 94% of the participants (*n*=29) in the intervention group completed, at least, the 80% of the intervention program sessions. There was a missing value for 20-m shuttle run, 4×10-m shuttle run, handgrip, standing long jump and prone bridging, and 2 missing values for 6-m gait speed test, otherwise, all the participants completed both assessments.

The main baseline characteristics of participants are shown in **Table 1**. There were no baseline differences between the groups in any tests.

**Table 1:** Participants baseline characteristics.

|  | CONTROL GROUP |  | INTERVENTION GROUP |  | *p-values* |
| --- | --- | --- | --- | --- | --- |
|  | (n=31) |  | (n=29) |  |  |
| Sex (female %) | 18 (58) |  | 19 (66) |  | 0.743 |
| Age (years) | 42.13 | 11.68 | 45.41 | 10.04 | 0.249 |
| Height (cm) | 167.19 | 10.17 | 165.44 | 7.74 | 0.458 |
| Weight (kg) | 74.36 | 15.42 | 74.12 | 19.36 | 0.958 |
| Body mass index (kg/m²) | 26.47 | 4.56 | 26.85 | 5.77 | 0.782 |
| Waist circumference (cm) | 87.58 | 12.10 | 86.96 | 15.39 | 0.862 |
| 20-m shuttle run (stage) | 3.05 | 2.09 | 2.84 | 1.92 | 0.691 |
| 2-km walk (s) | 1107.45 | 102.43 | 1076.93 | 91.71 | 0.230 |
| 6-min walk (m) | 678.51 | 58.55 | 680.56 | 54.64 | 0.889 |
| 6-m gait speed (s) | 2.40 | 0.22 | 2.44 | 0.24 | 0.448 |
| Timed up & go (s) | 5.74 | 0.63 | 5.67 | 0.62 | 0.664 |
| 4x10-m shuttle run (s) | 14.08 | 1.68 | 14.39 | 1.55 | 0.473 |
| Handgrip* (kg) | 32.75 | 10.23 | 31.53 | 10.24 | 0.645 |
| Standing long jump (cm) | 134.61 | 33.67 | 130.17 | 29.13 | 0.588 |
| 30-s sit to stand (rep) | 16.97 | 2.76 | 16.24 | 2.06 | 0.256 |
| Prone bridging (s) | 80.48 | 32.17 | 75.48 | 31.18 | 0.544 |

Results are presented as mean (SD) except for sex variable n (%). \*Handgrip right and left arm mean

Responsiveness of health-related field-based physical fitness tests are shown in **Table 2**. There were no differences pre-post tests in the control group; whereas in the intervention group, differences pre-post tests were observed in all tests (all *P*<0.01), except in waist circumference (*P*=0.091). Additional analyses were performed adjusted for sex and age and the results remained unchanged (data not shown). The 2-km walk, 6-min walk, 6-m gait speed, timed up & go, 30-s sit to stand and prone bridging tests showed a large effect size (*Cohen’s d*˃0.8). A moderate effect size was found in the 20-m shuttle run, 4×10-m shuttle run and standing long jump tests (*Cohen’s d*˃0.5-0.8). The weight, BMI and handgrip tests showed a trivial effect size (*Cohen’s d*<0.20). In absolute data, the difference in weight, BMI, waist circumference and handgrip test pre-post intervention were –0.94 kg, –0.5kg/m^2^, –0.84 cm and 1.3kg, respectively. Additional analyses were performed using the data of handgrip relative to corporal weight and the results remained unchanged (data not shown).

**Table 2.**
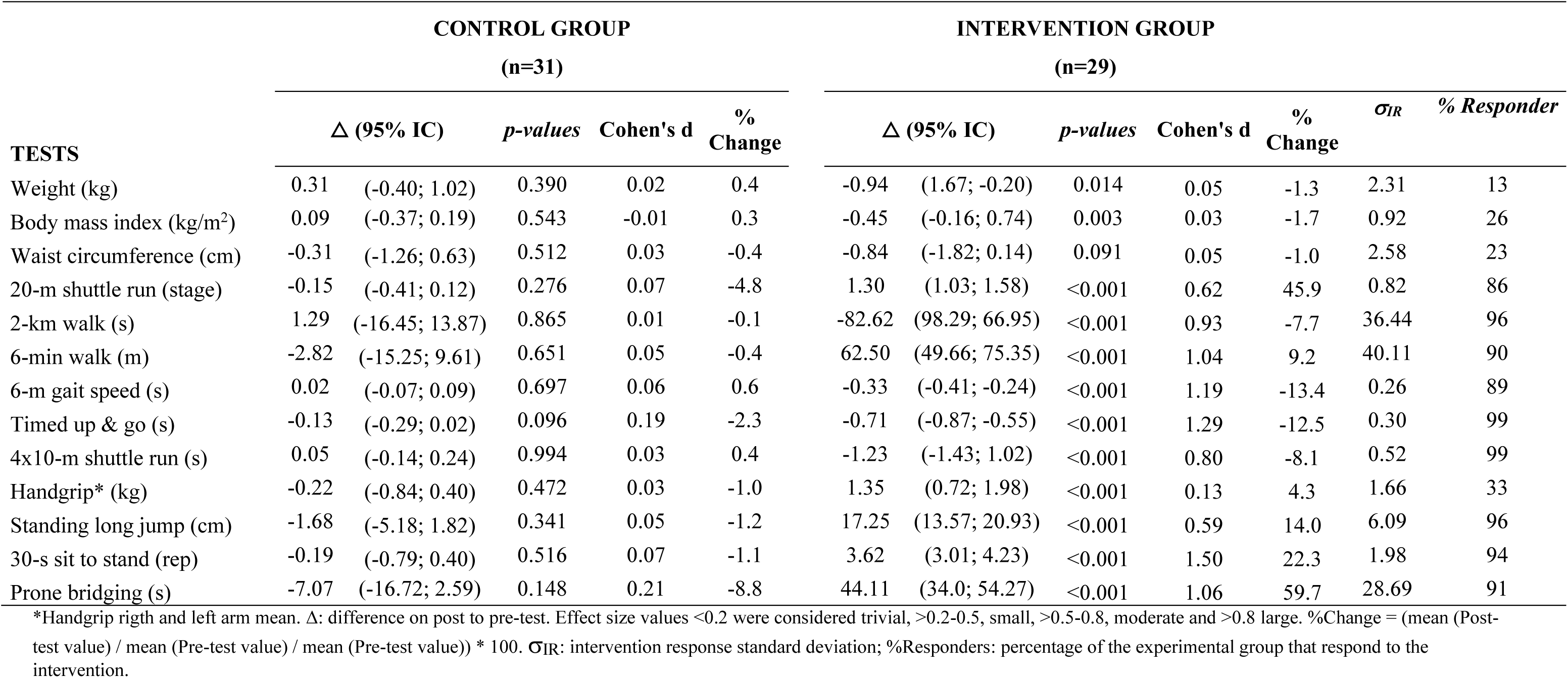
Pre-post differences for each groups and *percentage of the population that is expected to respond to the intervention*.

Figures 1 to 4 showed participants categorized into responders / no responders to intervention, based on individual change (Δ), 80%CI and SWC in all tests. Responder analysis showed a higher proportion of participants categorized as responders in the intervention group compared to the control group in all the variables analyzed. Among all of tests evaluated in the intervention group, we observed the highest proportion of individual responders in the 4×10-m shuttle run, 30-s sit-to-stand, 6-m gait speed and timed up & go tests (78%, 69%, 68 and 66% of responders, respectively) (figures 3-4). While in the 6-min walk, prone bridging, 2-km walk, 20-m shuttle run and standing long jump tests the proportion of individual responders were 57%, 54%, 52%, 46% and 46%, respectively, in the intervention group. The lower proportion of individual responders was observed in the weight, BMI, waist circumference and handgrip tests (<15%) in the intervention group. The proportion of responders in the control group ranged between 0 and 7% in all the tests studied, except in the timed up & go test with 23%.

**Figure 1.**
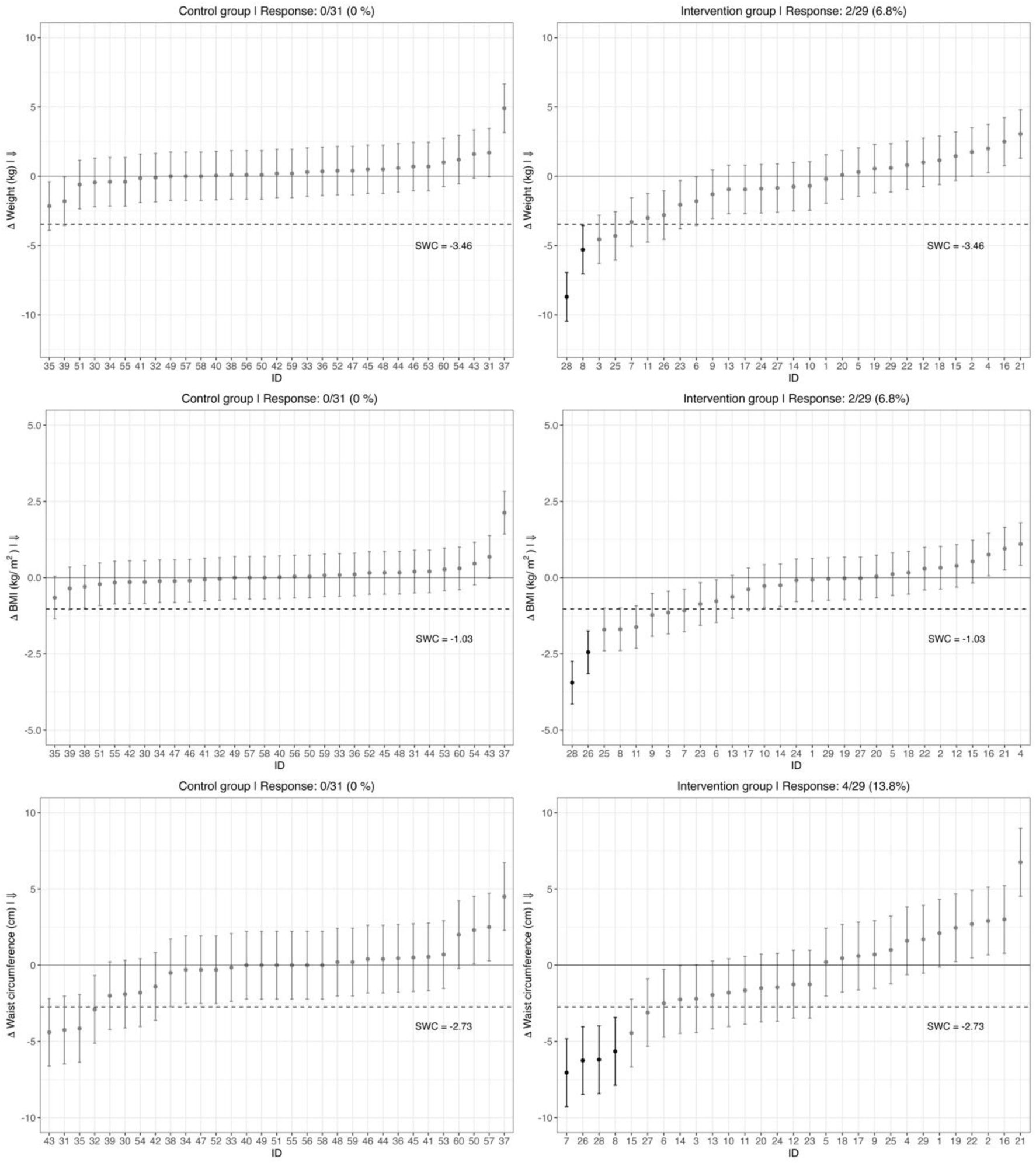
Participants categorized into responders / no responders to intervention in weight, BMI and waist circumference variables based on individual change (Δ), 80%CI and Smallest Worthwhile Change (SWC). Arrow next to y-axis title represents direction of the positive effect for each variable. Participants were classified as likely responders if the CI lies completely over or below the SWC (horizontal dotted line) depending on the interpretation of positive effect.

**Figure 2.**
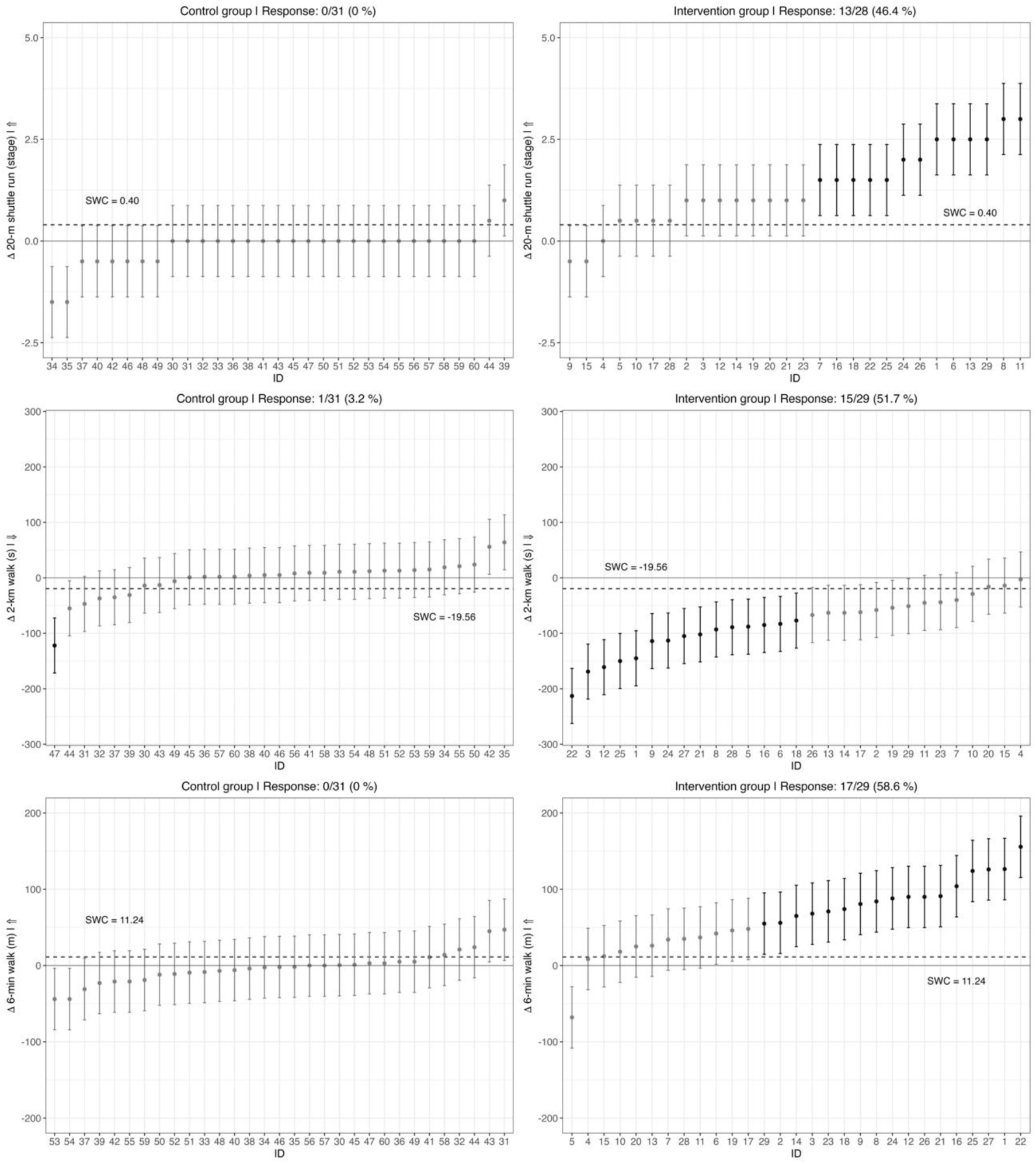
Participants categorized into responders / no responders to intervention in 20-m shuttle run, 2-km walk, and 6-min walk tests based on individual change (Δ), 80%CI and Smallest Worthwhile Change (SWC). Arrow next to y-axis title represents direction of the positive effect for each variable. Participants were classified as likely responder if the CI lies completely over or below the SWC (horizontal dotted line) depending of the interpretation of positive effect.

**Figure 3.**
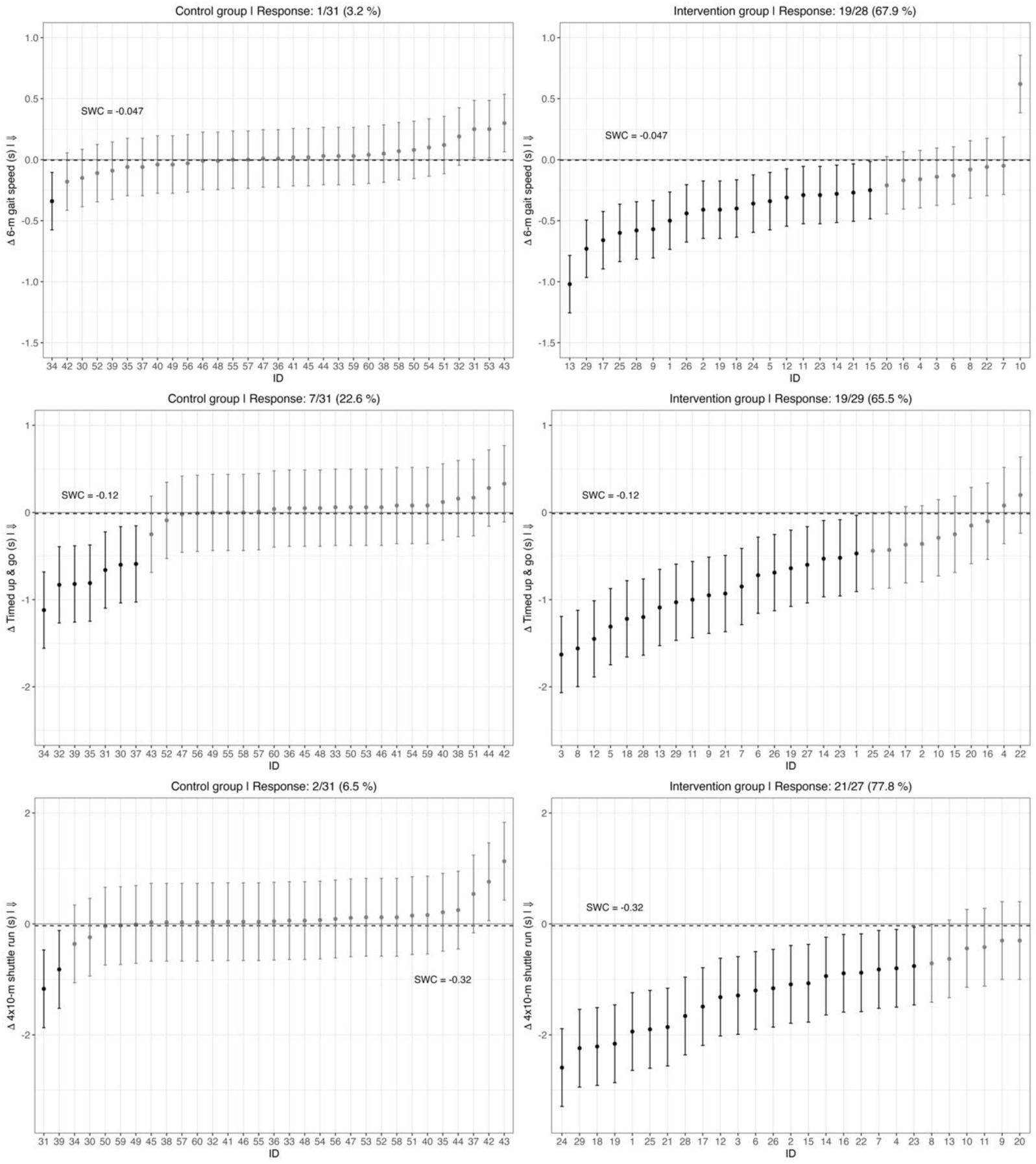
Participants categorized into responders / no responders to intervention in 6-m gait speed, timed up & go and 4×10-m shuttle run tests based on individual change (Δ), 80%CI and Smallest Worthwhile Change (SWC). Arrow next to y-axis title represents direction of the positive effect for each variable. Participants were classified as likely responder if the CI lies completely over or below the SWC (horizontal dotted line) depending on the interpretation of positive effect.

**Figure 4.**
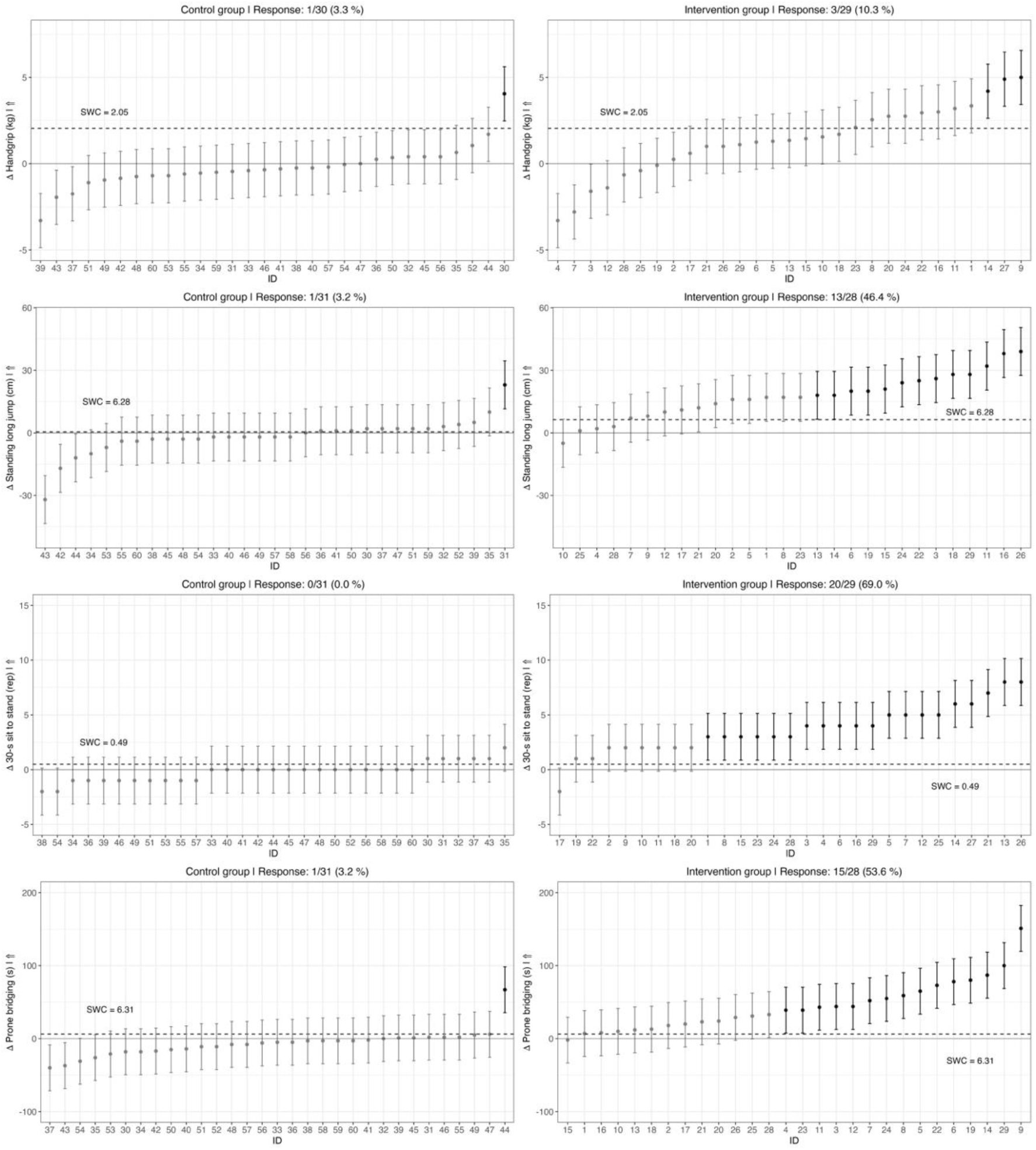
Participants categorized into responders / no responders to intervention in handgrip, standing long jump, 30-s sit-to-stand and prone bridging tests based on individual change (Δ), 80%CI and Smallest Worthwhile Change (SWC). Arrow next to y-axis title represents direction of the positive effect for each variable. Participants were classified as likely responder if the CI lies completely over or below the SWC (horizontal dotted line) depending of the interpretation of positive effect.

The percentage of the population that is expected to respond to the intervention is presented in **Table 2**. The expected proportion of responders based on σ_IR_ and SWC was higher to 85% for the 4×10-m shuttle run (99%), timed up & go (99%), 2-km walk (96%), standing long jump (96%), 30-s sit to stand (94%), prone bridging (91%), 6-min walk (90%), 6-m gait speed (89%) and 20-m shuttle run (86%) tests (table 2). While for the weight, BMI, waist circumference and handgrip tests were lower than 35%.

## Discussion

The aim of the present study was to determine the responsiveness of the health-related field-based physical fitness tests used in adults. Overall, all field-based physical fitness tests were found to be responsive after the intervention regardless sex and age with an effects size moderate to large, except for the weight, BMI and handgrip tests with trivial effect sizes. The highest proportion of individual responders was observed in the 4×10-m shuttle run, 30-s sit-to-stand, 6-m gait speed and timed up & go tests. The proportion of the population that is expected to respond to the intervention was higher that 85% in most of the health-related field-based fitness tests evaluated, but ≤ 33% in weight, BMI, waist circumference and handgrip tests. To the best of the authors’ knowledge, no previous studies have addressed the responsiveness on the health-related field-based physical fitness tests used in healthy adult population after an exercise intervention. Most of them have been focused on physical performance in clinical practice, in populations with pathologies and older adults with or without disease.

Responsiveness is considered the more essential propriety of an evaluative instruments, that is, the ability of a test to detect changes over time [28]. Therefore, apart from evaluating validity [3–7], reliability [9], feasibility and safety [10], the responsiveness needs also to be examined.

Our results displayed modest magnitude of change in whole-body weight (−0.94 kg), BMI (−0.50 kg/m^2^) and waist circumference (−0.84 cm) after exercise training in adults with a mean age of 45 years and a mean BMI of 26.9 kg/m^2^. The effects of moderate-intensity continuous training (MICT) combined or not with resistance training, HIIT or, MICT vs. HIIT on body composition has been analysed in recent years. It has been widely proven that exercise has a positive effect on body-weight, fat mass or waist circumference in adults, but small effects of change [33–37]. These studies concluded that the effect of exercise intervention might be influenced by age (18-65 years), complications (obesity), duration (>6 weeks), frequency, intensity, time efficiency and exercise modality. In addition, diet control is an important variable to the control of the body weight since a combination of diet with exercise has proved to be more effective than exercise alone regarding the reduction of body mass and fat mass [38].

Regarding field-based cardiorespiratory fitness tests, the results showed moderate and large magnitude of change after exercise training. At the individual level, ∼50-55% of the participants were responders. The proportion of the population that is expected to respond to the intervention was ≥86% in all tests. Responsiveness of 20-m shuttle run test has not been defined in healthy adult populations. A study developed in asthmatic adolescents showed an increase in the number of stages completed (7±1.4 to 10.1±1.3) in the 20-m shuttle run test after an individualized aerobic training program, and concluded that this test was sensible to change over time [39]. We observed a moderate magnitude of change after exercise training, with an improvement of 1.3 stages in non-active healthy adults. A study conducted in healthy non-athletic middle-aged adults observed a decrease in the time (minutes) spent to complete the 2-km walk test (13.1±1.0 to 15.1±0.6 in males; 15.1±1.0 to 16.3±1.1 in females) after 15-week walking training [40]. The authors concluded that the 2-km walk test can be used as a reasonably accurate field-based test to predict changes in the cardiorespiratory capacity due to aerobic training. These results are in line with our study in healthy non-active middle-aged adults, who improved 82 s in the 2-km walk test after the exercise training program. With respect to the 6-min walk test, it is commonly employed to evaluate responsiveness to rehabilitation or exercise program in population with chronic respiratory diseases, cardiovascular disease and older adults [41–43]. Available evidence suggested a minimally important difference of 30 meters in 6-min walk test in adults with chronic respiratory disease. Responsiveness data of 6-min walk test in healthy adults are limited. A study in adults in military service considered an improvement in more than 81.3 meters (via minimal detectable change (MDC)) in 6-min walk test as a true change in performance [44]; while our analyses showed an average increase of 62.5 meters after the intervention in non-active adults. Despite limited evidence in responsiveness of cardiorespiratory fitness tests in health adults, the 20-m shuttle run, 6-min and 2-km walk tests are responders to an exercise training program in or sample. The verbal encouragement and patient motivation are very important when evaluate a running or walking cardiorespiratory fitness tests influencing in the person’s effort capacity, especially in longer distance and self-regulated pace tests.

In field-based motor fitness tests, we observed a large magnitude of change in 6-m gait speed (−0.3) and timed up & go tests (−0.7 s), and moderate to the 4×10-m shutter run test (−1.2 s) after exercise training; and ∼66-78% of the participants were responders. The proportion of the population that is expected to respond to the intervention was ≥89% in all tests. Responsiveness of these tests has not been defined in healthy adult populations. A reduction of 1.0 s for the timed up & go test has been determined to be the MDC in older adults with type 2 diabetes [45], 0.5 s in people with epilepsy [46]. A study in older adults showed that the 4-m gait speed test is responsive with 0.05 m/s and 0.1 m/s indicating a substantial change (via small meaningful change) [47]. No responsiveness data has been reported in the 4×10-m shuttle run test. The 6-m gait speed and timed up & go tests are frequently used to evaluate motor fitness (i.e., speed of reaction, speed of movement or gait speed, agility, coordination) in older adults, while the 4×10-m shuttle run test is frequently used in the young population, making it difficult to find responsive data in adult population.

Regarding responsiveness of muscular strength tests, we observed significant but very small effects (*P* <0.001; *Cohen’s d* = 0.13) on handgrip test after exercise training, with a change of 1.3 kg. This is in line with previous studies in older adults, where change above 1.6 kg was considered a real change [48]. A recent metanalytical review examined the effects of different exercise training on handgrip strength in healthy community-dwelling older adults of 60 years or older. The effects change was small with a large effect in favor of program based on task-specific training and multimodal training modes to provide an appropriate stimulus to improve handgrip strength [49]; which is an interesting contribution to take into account when evaluating the efficacy of interventions with exercise training. Our intervention has incorporated different training focuses (cardiorespiratory, muscular and motor fitness (coordination, balance, agility and speed)), where grip strength was training thought grips and tractions with TRX (suspension training), elastic bands, dumbbell and kettlebell. However, initial level of handgrip strength of participants, intervention time (>3 months), intensity or task-specific training should be also taken into consideration to achieve a greater change.

The standing long jump responded after exercise training, with a moderate magnitude of change of 17.3 cm; the 46% of the participants were responders and the proportion of the population that is expected to respond to the intervention was ≥96%. Responsiveness of standing long jump test has not been defined in healthy adult populations. The 30-s chair stand and prone bridging tests showed a large magnitude of change after exercise training, the 69% and 54% of the participants were responders, respectively. The proportion of the population that is expected to respond to the intervention was ≥91% in both tests. A MDC value of 3.24 in adults with low back pain has been defined to the 30-s chair stand test; that is, an improvement in more than 3.24 repetitions has been considered a true change in performance [50]. Other studies have observed similar values in older adults with type 2 diabetes or dementia [45, 51]. Which is in line with our results, participants increased an average of 3.6 repetition more, after the exercise training intervention. Our analyses showed an average increase of ∼44 s in the prone bridging test after exercise training in healthy non-active adults. In a study developed in younger and older adults (20-35 and 60-79 years old, respectively), an improvement of 57 s in prone bridging test was considered a true change in performance (via MDC) [52]; this improvements in the prone bridging test is relatively higher that the change observed in our study, authors affirm that the sample was probably more fit and active than the general population. Thus, more studies about responsiveness of standing long jump, 30-s sit-to-stand and prone bridging tests in healthy adults are required.

## Strength and Limitations

This is the first study that analyses of responsiveness of a wide range of health-related field-based physical fitness tests, and after a thoroughly controlled a supervised training based on multimodal exercises during 12 weeks in healthy adults.

The responsiveness of a field-based physical fitness test is relevant in outcome studies as well as practical application in different settings (e.g.: clinical, sport club, etc). Additionally, the ratio administration time-evaluator-participant is more efficient than in laboratory clinical assessments.

Also, an exercise programme, based on the guidelines proposed by the American College of Sports Medicine for 12 weeks that includes 3 supervised weekly sessions of multicomponent exercise (i.e. including aerobic, muscle strengthening and motor fitness activities) with self-loading and low-cost equipment, shows positive results in improving physical fitness in adults.

The lack of control of factors that could affect the test performance, such as genetics, psychological issues, or/and diet control is the main limitation of this study

## Conclusions

Overall, most of the health-related field-based physical fitness tests analysed were found to be responsive after a multicomponent exercise training intervention regardless sex and age; except for changes in anthropometric outcomes and isometric handgrip strength, that were less responsive than the others and might require specifically targeted interventions to respond further. Assessing health-related physical fitness through field-based tests can be a useful assessment tool for determining the efficacy of different exercise program.

## Contributions to knowledge

What does this study add to existing knowledge?

- This is the first study based on scientific evidence to determine

What are the key implications for public health interventions, practice, or policy?

- This study underscores the critical importance of evaluating both feasibility and safety, delineating the essential components required for such assessments. These attributes—feasibility and safety—are indispensable criteria in the construction of a health-related, field-based physical fitness test battery for population-level assessment.

## Acknowledgments

We thank all participants and evaluators who took part in the ADULT-FIT study.

## Author’s contributions

MCG and JCP conceived the study idea. MCG led the writing of the review and carried out methodological procedures with CCL, SSP, JJI and JCP. All authors discussed the results and contributed to the final manuscript and agreed upon the order of presentation of the authors. All authors have read and approved the final manuscript.

## Funding

This project was supported by the Ministry of Economy, Industry and Competitiveness in the 2017 call for R&D Projects of the State Program for Research, Development and Innovation Targeting the Challenges of the Company; National Plan for Scientific and Technical Research and Innovation 2013–2016 (DEP2017-88043-R). National Plan for Scientific and Technical Research and Innovation 2017-2020 (PN / EPIF-FPU-CT / FPU20/02938), and the Regional Government of Andalusia and University of Cadiz: Research and Knowledge Transfer Fund (PPIT-FPI19-GJ4F-10).

## Data Availability

The authors declare that all relevant data are included in the article and/or its supplementary information files.

## Declarations

### Ethics Approval and Consent to Participate

All participants provide voluntarily written informed consent to be part of the present study. The study was approved by the Committee for Research of Cadiz, Spain.

## Consent for Publication

Not applicable.

## Competing interest

The authors declare that they have no financial interest directly or indirectly related to the work submitted for publication.

## Notes

### Competing Interest Statement

The authors have declared no competing interest.

### Author Declarations

The Cadiz Research Ethics Committee gave ethical approval for this work.

### Summary of Updates

The size of the images has been adjusted for better viewing of the manuscript.

## References

1. Kaminsky LA, Arena R, Ellingsen Ø, Harber MP, Myers J, Ozemek C, Ross R. Cardiorespiratory fitness and cardiovascular disease-the past, present, and future. Progress in cardiovascular diseases. 2019;62(2):86–93.

2. Kodama S, Saito K, Tanaka S, Maki M, Yachi Y, Asumi M, et al. Cardiorespiratory fitness as a quantitative predictor of all-cause mortality and cardiovascular events in healthy men and women: a meta-analysis. Jama. 2009;301(19):2024–35.

3. Garcia-Hermoso A, Cavero-Redondo I, Ramirez-Velez R, Ruiz JR, Ortega FB, Lee DC, Martinez-Vizcaino V. Muscular Strength as a Predictor of All-Cause Mortality in an Apparently Healthy Population: A Systematic Review and Meta-Analysis of Data From Approximately 2 Million Men and Women. Arch Phys Med Rehabil. 2018;99(10):2100–13 e5.

4. Harber M, Kaminsky L, Arena R, Blair S, Franklin B, Myers J, et al. Impact of cardiorespiratory fitness on all-cause and disease-specific mortality: advances since 2009. Progress in cardiovascular diseases. 2017;60:11–20.

5. Castro-Piñero J, Marin-Jimenez, N., Fernandez-Santos, J. R., Martin-Acosta, F., Segura-Jimenez, V., Izquierdo-Gomez, R., …, Cuenca-Garcia M. Criterion-Related Validity of Field-Based Fitness Tests in Adults: A Systematic Review. Journal of clinical medicine. 2021;10:3743.

6. Marin-Jimenez N, Cruz-Leon C, Sanchez-Oliva D, Jimenez-Iglesias J, Caraballo I, Padilla-Moledo C, Castro-Piñero J. Criterion-Related Validity of Field-Based Methods and Equations for Body Composition Estimation in Adults: A Systematic Review. Current Obesity Reports. 2022:1–14.

7. Marín-Jiménez N, Cruz-León C, Perez-Bey A, Conde-Caveda J, Grao-Cruces A, Aparicio VA, …, Cuenca-García M. Predictive Validity of Motor Fitness and Flexibility Tests in Adults and Older Adults: A Systematic Review. Journal of Clinical Medicine. 2022;11:328.

8. Soysal P, Hurst C, Demurtas J, Firth J, Howden R, Yang L, et al. Handgrip strength and health outcomes: Umbrella review of systematic reviews with meta-analyses of observational studies. Journal of Sport and Health Science. 2021;10:290–5.

9. Cuenca-Garcia M, Marin-Jimenez N, Perez-Bey A, Sánchez-Oliva D, Camiletti-Moiron D, Alvarez-Gallardo IC, Castro-Piñero J. Reliability of Field-Based Fitness Tests in Adults: A Systematic Review. Sports Medicine. 2022:1–20.

10. Cruz-León C, Marín-Jiménez N, Sánchez-Parente S, Borges Cosic M, Grao-Cruces A, Castro-Piñero J, Cuenca-Garcia M. Feasibility and Safety in health-related field-based physical fitness tests in adult population: The ADULT-FIT project. International Journal of Behavioral Nutrition and Physical Activity. 2025; Under Rev.

11. Terwee CB, Bot SD, de Boer MR, van der Windt DA, Knol DL, Dekker J, et al. Quality criteria were proposed for measurement properties of health status questionnaires. Journal of clinical epidemiology. 2007:34–42.

12. Marfell-Jones M, Olds T, Stewart A, Carter L. ISAK accreditation handbook 2006.

13. Leger LA, Mercier D, Gadoury C, Lambert J. The multistage 20 metre shuttle run test for aerobic fitness. Journal of sports sciences. 1988;6:93–101.

14. Oja P, Laukkanen R, Pasanen M, Tyry T, Vuori I. A 2-km walking test for assessing the cardiorespiratory fitness of healthy adults. 1991;12:356–62.

15. Mänttäri A, Suni J, Sievänen H, Husu P, Vähä-Ypyä H, Valkeinen H, et al. Six-minute walk test: a tool for predicting maximal aerobic power (VO 2 max) in healthy adults. Clinical physiology and functional imaging. 2018;38(6):1038–45.

16. Rikli RE, Jones CJ. Development and validation of a functional fitness test for community-residing older adults. Journal of aging and physical activity. 1999;7:129–61.

17. Woo J, Yau F, Leung J, Chan R. Peak oxygen uptake, six-minute walk distance, six-meter walk speed, and pulse pressure as predictors of seven year all-cause and cardiovascular mortality in community-living older adults. Experimental Gerontology. 2019;124: 110645.

18. Ruiz JR, Castro-Piñero J, España-Romero V, Artero EG, Ortega FB, Cuenca MM, et al. Field-based fitness assessment in young people: the ALPHA health-related fitness test battery for children and adolescents. British journal of sports medicine. 2011;45(6):518–24.

19. Ruiz-Ruiz J, Mesa JL, Gutiérrez A, Castillo MJ. Hand size influences optimal grip span in women but not in men. The Journal of hand surgery. 2002;27(5):897–901.

20. Tong TK, Wu S, Nie J. Sport-specific endurance plank test for evaluation of global core muscle function. Physical Therapy in Sport. 2014;15:58–63.

21. Edwards JJ, Griffiths M, Deenmamode AH, O’Driscoll JM. High-Intensity Interval Training and Cardiometabolic Health in the General Population: A Systematic Review and Meta-Analysis of Randomised Controlled Trials. Sports Med. 2023:1–11.

22. Lopez P, Radaelli R, Taaffe DR, Newton RU, Galvão DA, Trajano GS, Pinto RS. Resistance training load effects on muscle hypertrophy and strength gain: Systematic review and network meta-analysis. Medicine and science in sports and exercise. 2021;53:1206.

23. Markov AHL, Chaabene H. Effects of Concurrent Strength and Endurance Training on Measures of Physical Fitness in Healthy Middle-Aged and Older Adults: A Systematic Review with Meta-Analysis. Sports Med. 2023;53:437–55.

24. Liguori G, American College of Sports M. ACSM’s guidelines for exercise testing and prescription. USA 2021.

25. Borg GA. Psychophysical bases of perceived exertion. Medicine and science in sports and exercise. 1982;14(5):377–81.

26. El-Amrawy F, Nounou MI. Are currently available wearable devices for activity tracking and heart rate monitoring accurate, precise, and medically beneficial? Healthcare informatics research. 2015;21(4):315–20.

27. Cohen J. Statistical power analysis for the behavioral sciences: Routledge; 2013.

28. Hopkins WG. A Spreadsheet for Monitoring an Individual’s Changes and Trend. Sportscience. 2017;21.

29. Bonafiglia JT, Nelms MW, Preobrazenski N, LeBlanc C, Robins L, Lu S, et al. Moving beyond threshold-based dichotomous classification to improve the accuracy in classifying non-responders. Physiological reports. 2018;6:e13928.

30. Hopkins WG. Individual responses made easy. American Physiological Society Bethesda, MD; 2015. p. 1444–6.

31. Swinton PA, Hemingway BS, Saunders B, Gualano B, Dolan E. A statistical framework to interpret individual response to intervention: paving the way for personalized nutrition and exercise prescription. Frontiers in nutrition. 2018;5:360939.

32. Team RC. R: A language and environment for statistical computing. R Foundation for Statistical Computing, Vienna, Austria. http://www.R-project.org/. 2016.

33. Guo Z, Li M, Cai J, Gong W, Liu Y, Liu Z. Effect of high-intensity interval training vs. moderate-intensity continuous training on fat loss and cardiorespiratory fitness in the young and middle-aged a systematic review and meta-analysis. International Journal of Environmental Research and Public Health. 2023;20(6):4741.

34. Keating SE, Johnson NA, Mielke GI, Coombes JS. A systematic review and meta-analysis of interval training versus moderate-intensity continuous training on body adiposity. Obesity reviews. 2017;18(8):943–64.

35. Wewege M, Van Den Berg R, Ward R, Keech A. The effects of high-intensity interval training vs. moderate-intensity continuous training on body composition in overweight and obese adults: a systematic review and meta-analysis. Obesity reviews. 2017;18(6):635–46.

36. Wilson JM, Marin PJ, Rhea MR, Wilson SM, Loenneke JP, Anderson JC. Concurrent training: a meta-analysis examining interference of aerobic and resistance exercises. The Journal of Strength & Conditioning Research. 2012;26(8):2293–307.

37. Sultana RN, Sabag A, Keating SE, Johnson NA. The effect of low-volume high-intensity interval training on body composition and cardiorespiratory fitness: a systematic review and meta-analysis. Sports Medicine. 2019;49:1687–721.

38. Clark JE. Diet, exercise or diet with exercise: comparing the effectiveness of treatment options for weight-loss and changes in fitness for adults (18–65 years old) who are overfat, or obese; systematic review and meta-analysis. Journal of Diabetes & Metabolic Disorders. 2015;14:1–28.

39. Ahmaidi SB, Varray AL, Savy-Pacaux AM, Pnefaut CG. Cardiorespiratory fitness evaluation by the shuttle test in asthmatic subjects during aerobic training. Chest. 1993;103(4):1135–41.

40. Laukkanen RMT, Kukkonen-Harjula TK, Oja P, Pasanen ME, Vuori IM. Prediction of change in maximal aerobic power by the 2-km walk test after walking training in middle-aged adults. International journal of sports medicine. 2000;21:113–6.

41. Holland AE, Spruit MA, Troosters T, Puhan MA, Pepin V, Saey D, … & Singh SJ. An official European Respiratory Society/American Thoracic Society technical standard: field walking tests in chronic respiratory disease. European Respiratory Journal. 2014;44:1428–46.

42. O’Keeffe S, Lye M, Donnellan C, Carmichael D. Reproducibility and responsiveness of quality of life assessment and six minute walk test in elderly heart failure patients. Heart. 1998;80:337.

43. Bellet RN, Adams L, Morris NR. The 6-minute walk test in outpatient cardiac rehabilitation: validity, reliability and responsiveness: a systematic review. Physiotherapy. 2012;98:277–86.

44. Marín-Jiménez N, Sánchez-Parente S, Expósito-Carrillo P, Jiménez-Iglesias J, Álvarez-Gallardo IC, Cuenca-García M, & Castro-Piñero J. Criterion-related validity and reliability of the 2-km walk test and the 20-m shuttle run test in adults: The role of sex, age and physical activity level. Journal of Science and Medicine in Sport. 2023;26:267–76.

45. Alfonso-Rosa RM, del Pozo-Cruz B, del Pozo-Cruz J, Sanudo B, & Rogers ME. Test–retest reliability and minimal detectable change scores for fitness assessment in older adults with type 2 diabetes. Rehabilitation Nursing Journal. 2014;39:260–8.

46. Aktar B, Balci B, Oztura I, & Baklan B. The test-retest reliability and minimal detectable change of the six-minute walk test, timed up and go test, and 30-second chair stand test in people with epilepsy. Physiotherapy theory and practice. 2023:1–10.

47. Perera S, Mody SH, Woodman RC, & Studenski SA. Meaningful change and responsiveness in common physical performance measures in older adults. Journal of the American Geriatrics Society. 2006;54:743–9.

48. Bohannon R. Test-Retest Reliability of Measurements of Hand-Grip Strength Obtained by Dynamometry from Older Adults: A Systematic Review of Research in the PubMed Database. The Journal of frailty & aging. 2017;6(2):83–7.

49. Labott B, H. B, Morat M, Morat T, Donath L. Effects of Exercise Training on Handgrip Strength in Older Adults: A Meta-Analytical Review. Gerontology. 2019;65:686–98.

50. Kahraman T, Kahraman BO, Sengul YS, & Kalemci O. Assessment of sit-to-stand movement in nonspecific low back pain: a comparison study for psychometric properties of field-based and laboratory-based methods. International Journal of Rehabilitation Research. 2016;39:165–70.

51. Blankevoort CG, Van Heuvelen MJ, & Scherder EJ. Reliability of six physical performance tests in older people with dementia. Physical therapy. 2013;93(1):69–78.

52. Bohannon RW, Steffl M, Glenney SS, Green M, Cashwell L, Prajerova K, & Bunn J. The prone bridge test: Performance, validity, and reliability among older and younger adults. Journal of bodywork and movement therapies. 2018;22(2):385–9.

